# The effect of reactive balance training on falls in daily life: an updated systematic review and meta-analysis

**DOI:** 10.1101/2022.01.27.22269969

**Authors:** Augustine Joshua Devasahayam, Kyle Farwell, Bohyung Lim, Abigail Morton, Natalie Fleming, David Jagroop, Raabeae Aryan, Tyler Mitchell Saumur, Avril Mansfield

## Abstract

**Objective:** Reactive balance training is an emerging approach to reduce falls risk in people with balance impairments. The purpose of this study was to determine the effect of reactive balance training on falls in daily life among individuals at increased risk of falls, and to document associated adverse events.

**Data sources:** Databases searched were Ovid MEDLINE (1946-November 2020), Embase Classic and Embase (1947-November 2020), Cochrane Central Register of Controlled Trials (2014-November 2020), Physiotherapy Evidence Database (PEDro; searched on 9 November 2020).

**Study selection:** Randomized controlled trials of reactive balance training were included. The literature search was limited to English language. Records were screened by two investigators separately.

**Data extraction:** Outcome measures were number of participants who reported falls after training, number of falls reported after training, and the nature, frequency, and severity of adverse events. Authors of included studies were contacted to obtain additional information.

**Data synthesis:** Twenty-five trials were included, of which 14 reported falls and 19 monitored adverse events. Participants assigned to reactive balance training groups were less likely to fall compared to control groups (fall risk ratio: 0.75, 95% confidence interval=[0.60, 0.92]; p=0.006, I^2^=37%) and reported fewer falls than control groups (rate ratio: 0.60, 95% confidence interval=[0.42, 0.86]; p=0.005, I^2^=83%). Prevalence of adverse events was higher in reactive balance training (29%) compared to control groups (19%; p=0.018).

**Conclusion:** RBT reduces the likelihood of falls in daily life for older adults and people with balance impairments. More adverse events were reported for reactive balance training than control groups.

**Impact:** Balance training that evokes balance reactions can reduce falls among people at increased risk of falls.

## INTRODUCTION

Falls are a common cause of injuries and deaths among people with balance impairments, such as older adults.^1^ Fall-related injuries result in significant healthcare costs to older adults and pose an economic burden to society.^2^ Exercise that includes balance training can reduce the rate of falls by about 25%.^3^ However, individualized exercise approaches seem to be more effective in reducing falls risk than others (e.g., based on the frequency, intensity, type, and duration of training).^3^

Effective balance control involves the ability to sustain postures, move between postures, and to react to perturbations (loss of balance).^4^ A fall happens when a person fails to respond to a balance perturbation,^5^ therefore, impaired balance reactions may increase the risk for falls. Indeed, a meta-analysis of 61 studies including 9,536 older adults reported that impaired reactive stepping was a significant risk factor for falls among older adults.^6^ ‘Conventional’ balance exercises mostly involve maintaining balance during static postures or during movement.^3^ Reactive balance training (RBT) is a type of balance training where participants repeatedly experience balance perturbations, and execute balance reactions to prevent a fall.^7^ RBT has been shown to improve control of balance reactions, and therefore, might help individuals respond to a loss of balance and prevent a fall, in daily life.^8-10^ Indeed, we previously reported in a meta-analysis of small randomized controlled trials that RBT reduces the rate of falls in daily life by almost half among people with increased fall risk, compared to other types of exercise or no intervention.^7^ This previous review was completed in 2015;^7^ more randomized controlled trials on RBT have been published since then.

The purpose of this study was to determine the effect of RBT, compared to either no or an active non-RBT control intervention, on falls in daily life in people who are at an increased risk of falls. Clinical interventions must balance intervention efficacy and harms.^11^ Therefore, our secondary aim was to document the nature, frequency, and severity of adverse events due to RBT, and to determine if there is an increased prevalence of adverse events with RBT compared to other types of exercise.

## METHODS

### Study design

This study is an update of a previous systematic review and meta-analysis,^7^ registered with PROSPERO database (CRD42020220552),^12^ conducted according to the Cochrane Handbook for Systematic Reviews of Interventions,^13^ and reported using the PRISMA statement for reporting systematic reviews and meta-analyses of randomized controlled studies.^14^

### Eligibility criteria

Studies that met the following criteria were included in this review: (1) published in English; (2) an experimental investigation of RBT; (3) included people at increased risk of falls due to impaired balance control, such as apparently healthy older adults (60 years or older), people with neurological conditions (e.g., stroke or Parkinson’s disease), people with COPD, people with lower-extremity amputations or joint replacement, or any other condition that increases the risk for falling; (4) included a control group that did not complete RBT; (5) randomly allocated participants to RBT and control groups; and (6) reported information about falls in daily life after the intervention and/or adverse events. RBT was defined as a training method where participants intentionally experience repeated loss of balance, with a goal of evoking balance reactions such as stepping or reach-to-grasp responses, so that participants can practice and improve control of balance reactions. The loss of balance can either be caused by an external force (e.g., a moving platform, push or pull from a therapist) or by the participant’s inability to maintain balance during voluntary movement.

### Data sources and search strategy

The literature search was conducted in MEDLINE® ALL (in Ovid, including Epub Ahead of Print, In-Process & Other Non-Indexed Citations, Ovid MEDLINE® Daily; 1946 to 5 November 2020), Embase (in Ovid, including Embase Classic; 1947 to 10 November 2020), Physiotherapy Evidence Database (PEDro; searched on 9 November 2020), and Cochrane Central Register of Controlled Trials (Ovid; 2014 to 10 November 2020) databases by an information specialist. Since the terminology related to RBT (also referred to as perturbation-based balance training) did not have standardized keywords or Medical Subject Headings (MeSH) terms, a search strategy involving multiple relevant keywords was used. A sample search strategy is provided as a supplementary material (Appendix A).

### Selection process

The titles of articles retrieved from the databases were screened by two reviewers independently and clearly ineligible titles were removed. The abstracts of the remaining articles were screened again by two reviewers independently to determine eligibility for this review. The full text of the article was read by the reviewers if they were unable to determine eligibility from the abstract. Disagreements between reviewers regarding inclusion of a study were resolved via discussion, or consultation with the study team if necessary. The reference lists of relevant review articles were also screened by the two reviewers using steps mentioned above to identify studies that might be eligible for this review. Authors of papers, including study protocols that met all but the last inclusion criterion (reported data on falls in daily life and/or adverse events), were contacted to determine if these data existed. Bibliographic references were managed using Endnote (version X5, Thomson Reuters, Philadelphia, Pennsylvania, USA).

### Data collection process

The following data were extracted from the articles selected for this review: details of the population studied; details of the RBT and control interventions; number of participants allocated to RBT and control groups; falls monitoring duration; number of participants in each group reporting one or more falls after completing the interventions; total number of falls per group reported after the interventions; and the nature, frequency, and severity of adverse events reported in RBT and control groups. The trial protocols and secondary publications were accessed to obtain the required information, if necessary and available. The corresponding authors of included studies were also contacted for additional details and to obtain any missing information. The data were extracted and compiled into a Microsoft Excel® spreadsheet. The three outcomes of interest from studies included in this review were: (1) number of participants who experienced one or more falls after the end of the intervention; (2) number of falls in daily life after the end of the intervention; and (3) nature, frequency, and severity of adverse events during the intervention. Adverse events were deemed to be severe if they were life threatening or resulted in permanent harm, moderate if they required hospitalization or resulted in withdrawal from the intervention (such that the participant no longer received the benefits of the intervention), or mild otherwise.

### Methodological quality and risk of bias assessment

Studies included in this review were rated for methodological and reporting quality by two reviewers independently using the PEDro scale.^15^ Disagreement during data extraction and quality assessment of the included articles were resolved by consensus after discussion between reviewers. If an agreement was not reached between two reviewers, the final scores were assigned by an independent third reviewer. The final PEDro scores were classified as good for ≥ 6, fair for 4 to 5, and poor for ≤ 3, based on previous sensitivity analyses with cut-offs set at 4 and 6.^15, 16^

Risk of bias was evaluated independently by two reviewers for all studies that assessed risk of falls using the revised Cochrane Risk of Bias Tool in five domains: (1) risk of bias arising from the randomization process; (2) risk of bias due to deviations from the intended interventions; (3) missing outcome data; (4) risk of bias in measurement of the outcome; and (5) risk of bias in selection of the reported results.^17, 18^ The studies were rated as: ‘high risk’ of bias if at least one out of five domains was at a high risk of bias or if there were ‘some concerns’ of bias for multiple domains; rated as ‘some concerns’ if at least one out of five domains had ‘some concerns’ of bias with no other domains at ‘high risk’ of bias; and rated as ‘low risk’ of bias if all five domains were at ‘low risk’ of bias.^17, 18^ Disagreement between reviewers during the risk of bias assessment was resolved by consensus after discussion, or by an independent third reviewer if consensus could not be reached.

### Data analysis

Data extracted from included studies were analysed using Review Manager (RevMan) software (version 5.4.1, The Cochrane Collaboration, Copenhagen, Denmark). Heterogeneity was assessed using chi-squared test of heterogeneity and I^2^ statistic.^19^ Data from included studies were pooled using random effects modeling when heterogeneity was significant (>30%).^13^ A subgroup meta-analysis was performed that excluded studies with a no-intervention control group. The effects of RBT on falls in daily life compared to control interventions were reported as fall risk ratios and fall rate ratios, where the fall risk ratio compared the number of participants who experienced one or more falls between groups and the fall rate ratio compared the number of falls between groups. Ninety-five percent confidence intervals (95% CIs) were calculated for the overall effects and the significance level was set at 0.05 for all analyses. The total number of participants with adverse events during RBT and control group training were summarized in percentages and compared using Fisher’s Exact test through SAS® software (SAS Institute, Inc., Cary, North Carolina).

### Reporting bias assessment

Funnel plot asymmetry was used to examine bias in the results of the meta-analyses of risk of falls and rate of falls as per Sterne *et al*.,^20^ recommendations. Two independent reviewers evaluated the possible sources of asymmetry on funnel plots due to selective non-reporting or non-publication of findings from the studies, poor methodological quality in studies with inflated effects, true heterogeneity between studies (e.g., due to difference in intensity of interventions), sampling variation, and random chance.^21^ Disagreement between reviewers during the reporting bias assessment was resolved by consensus after discussion or by an independent third reviewer.

### Certainty assessment

Certainty of evidence from studies that contributed data to the meta-analysis of risk of falls was assessed separately by two reviewers using five ‘Grading of Recommendations, Assessment, Development, and Evaluation (GRADE)’ domains (risk of bias, inconsistency, indirectness, imprecision, other considerations).^22^ The labels from GRADE certainty of evidence assessment, i.e., not serious, serious, and very serious, were used to assess the body of evidence we evaluated through meta-analysis.^23-25^ These labels correspond to high, moderate, and low certainty of evidence for randomized-controlled studies.^23-25^ The Cochrane Handbook for Systematic Reviews of Interventions was used to conduct the certainty assessment and prepare the summary of findings table through GRADEpro GDT software (GRADEpro guideline development tool, McMaster University and Evidence Prime Inc., Hamilton, Ontario, Canada).^26^ Disagreement between the reviewers during certainty assessment was resolved by consensus after discussion or by an independent third reviewer.

## RESULTS

### Study selection

For the purposes of this review, a ‘record’ refers to a document indexed in a database, a ‘report’ refers to a document supplying information about a study (e.g., paper or abstract), and a ‘study’ refers to an investigation of one or more interventions in a group of participants;^50^ a study can have several reports (e.g., protocol paper, primary publication, secondary publication(s)). A total of 5,606 records were identified through database searching and an additional 182 records were identified from other sources. The records included in our previous review^7^ were also screened (n=9).

After removing duplicates, the remaining records (n=3,491) were screened for eligibility through a title search. The abstracts of the remaining reports (n=518) were assessed for eligibility. One study included in the previous review was removed from this review due to pseudo-random group allocation.^51^ Another study from the previous review was an internal pilot study for a larger randomized controlled trial that was included in the current review;^52^ to avoid including duplicate data in the analysis, data from the pilot study were not included in the current review. After screening, 40 reports of 25 studies were included in this review (Figure 1). All reports for each included study are listed in Appendix B. The characteristics of the studies included in this review are presented in Table 1.

**Table 1:**
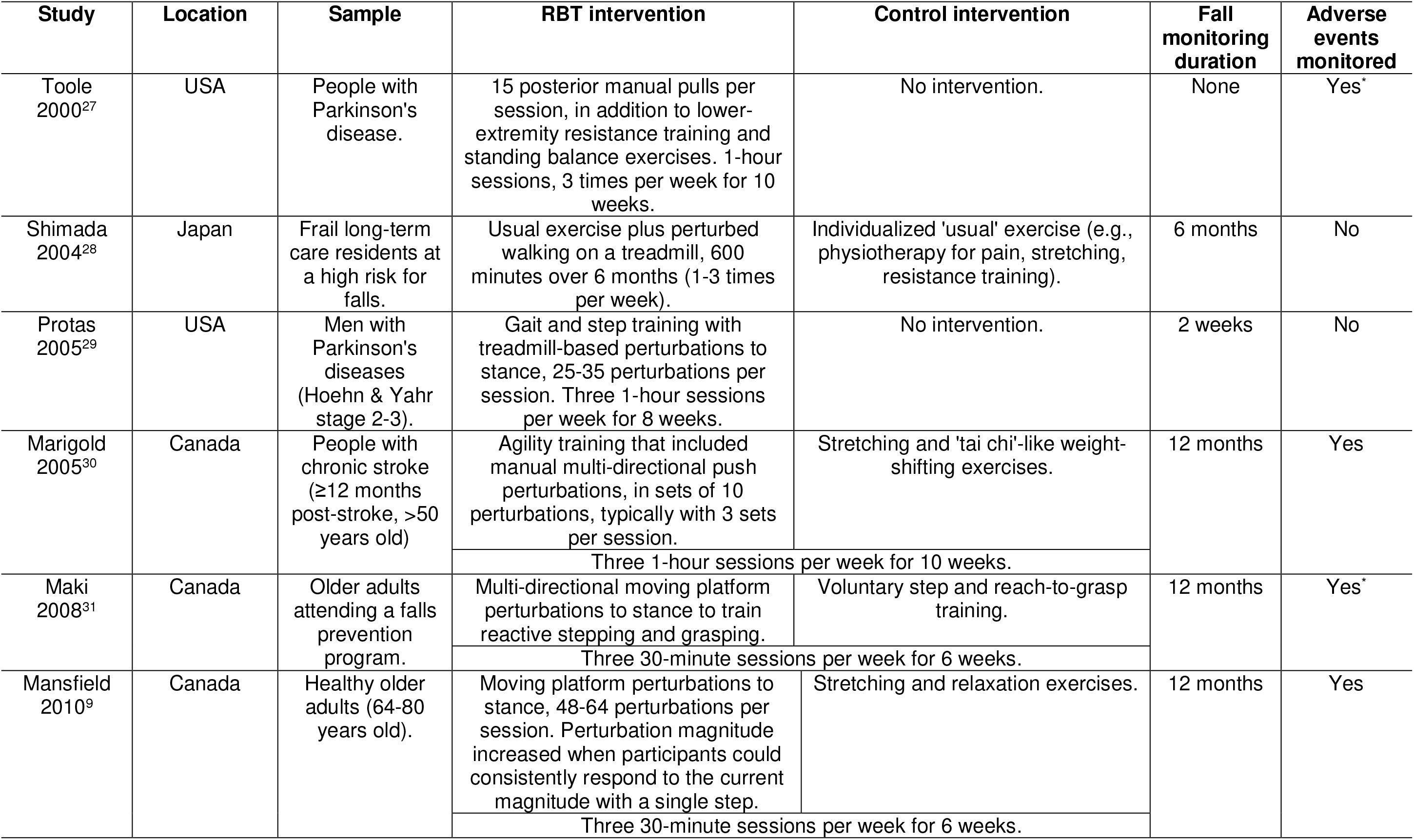
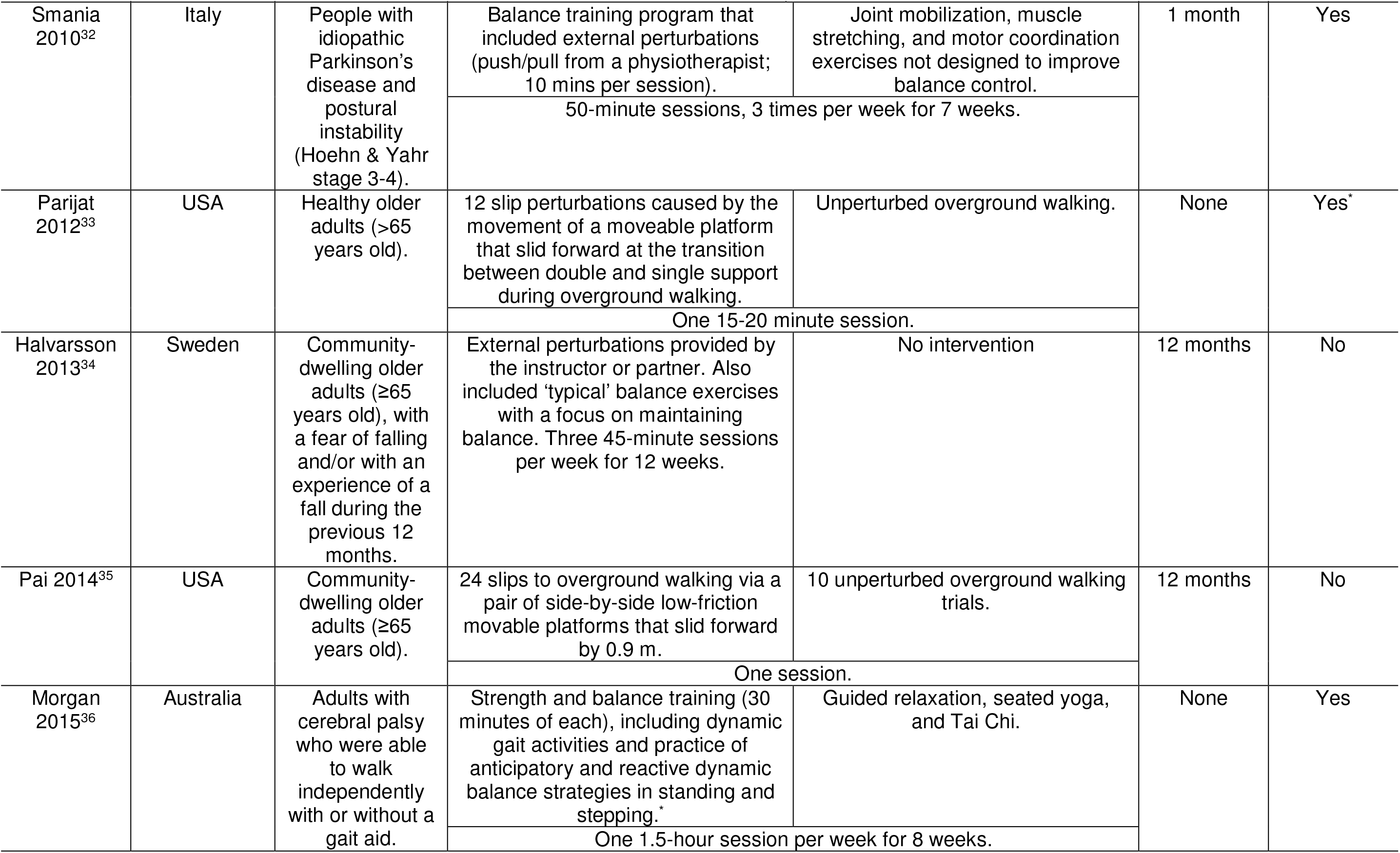
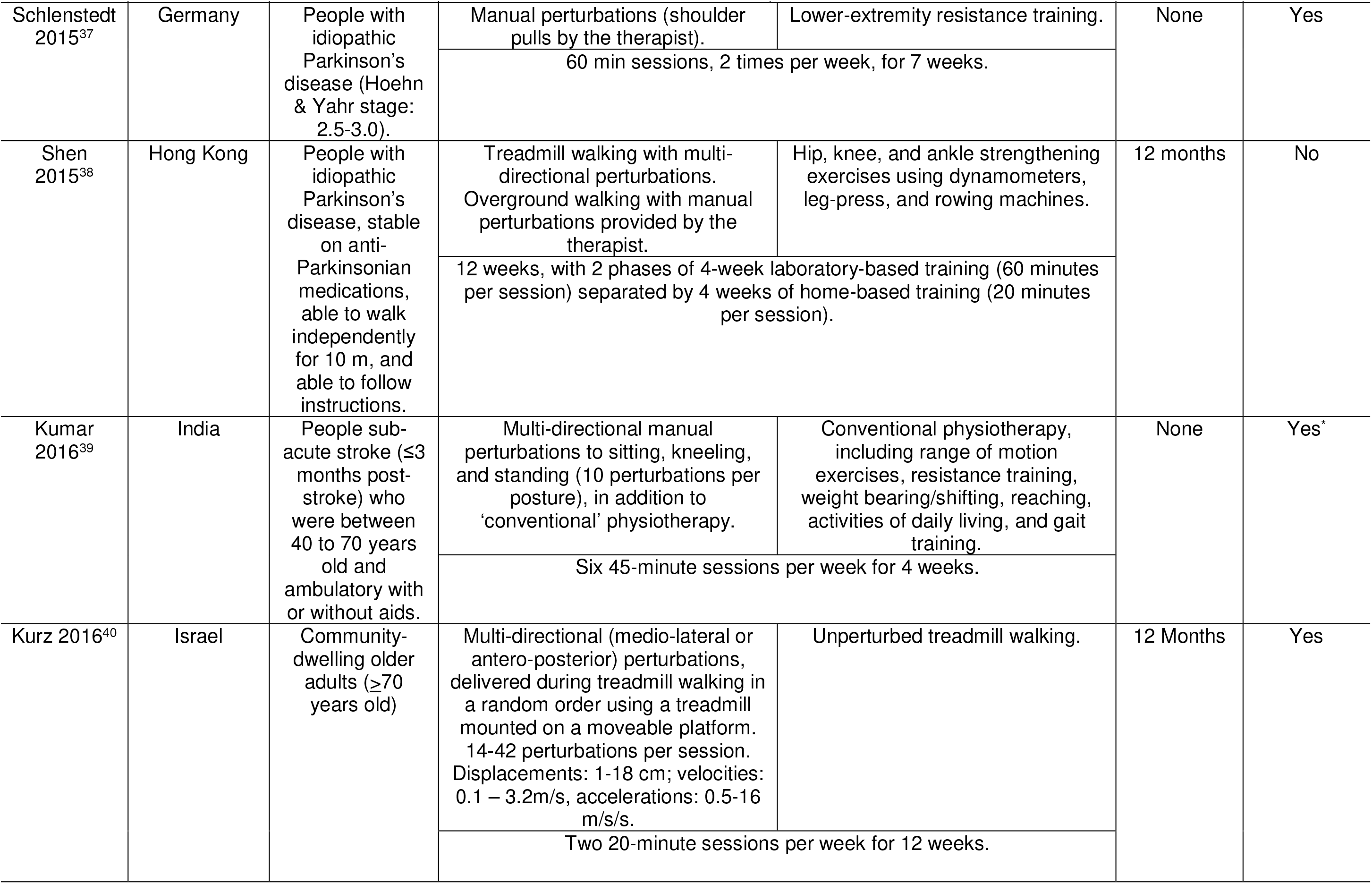
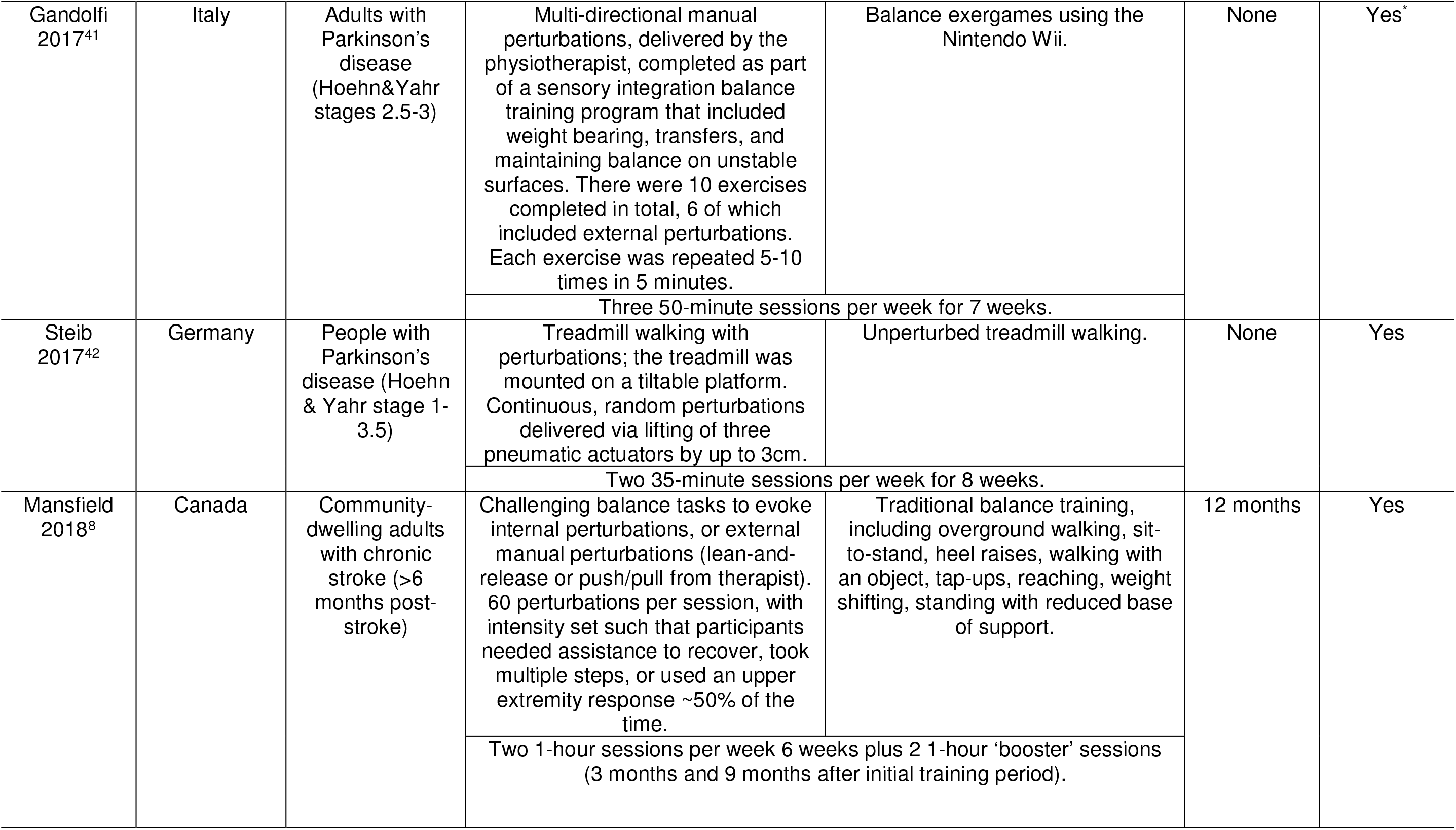
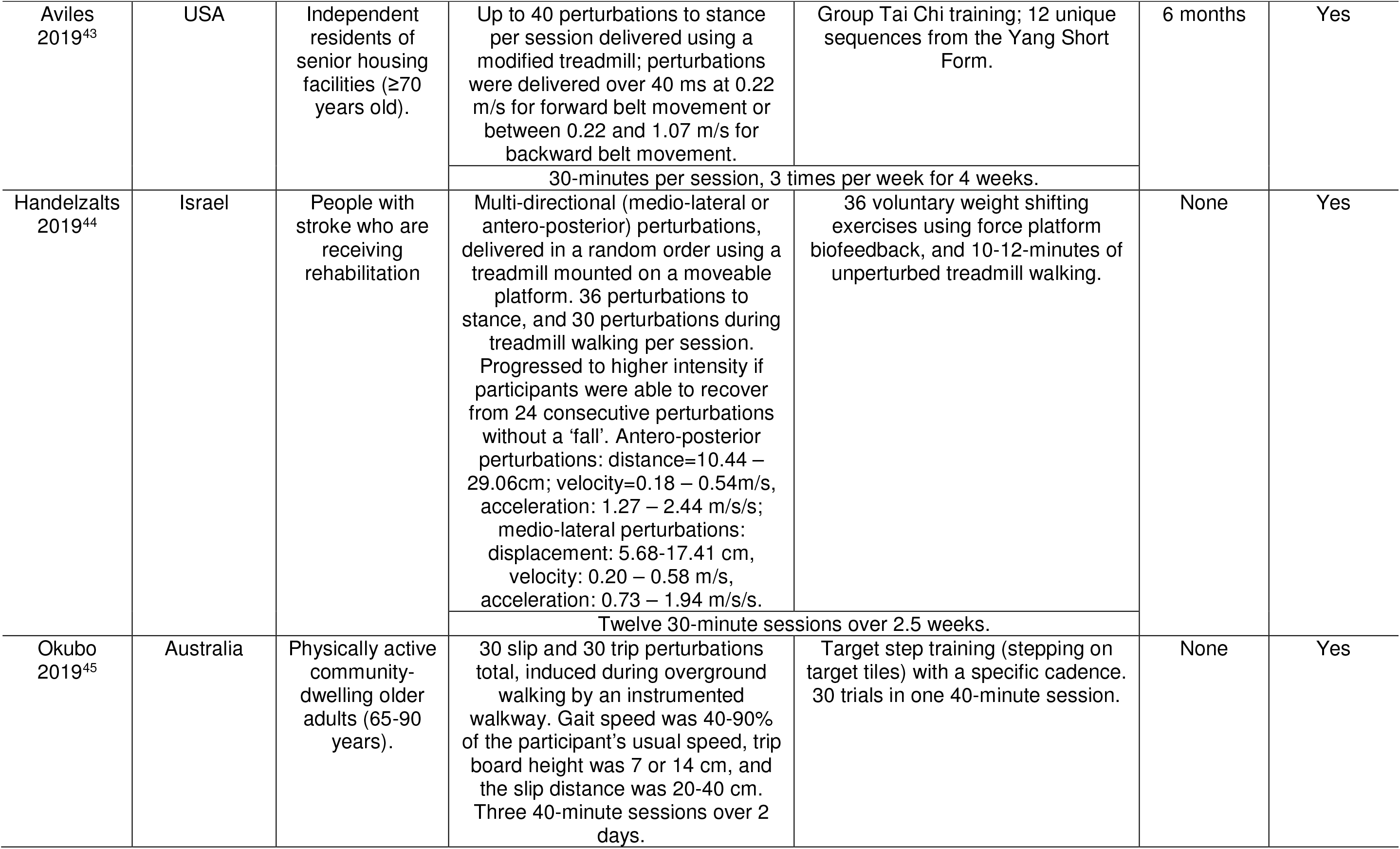
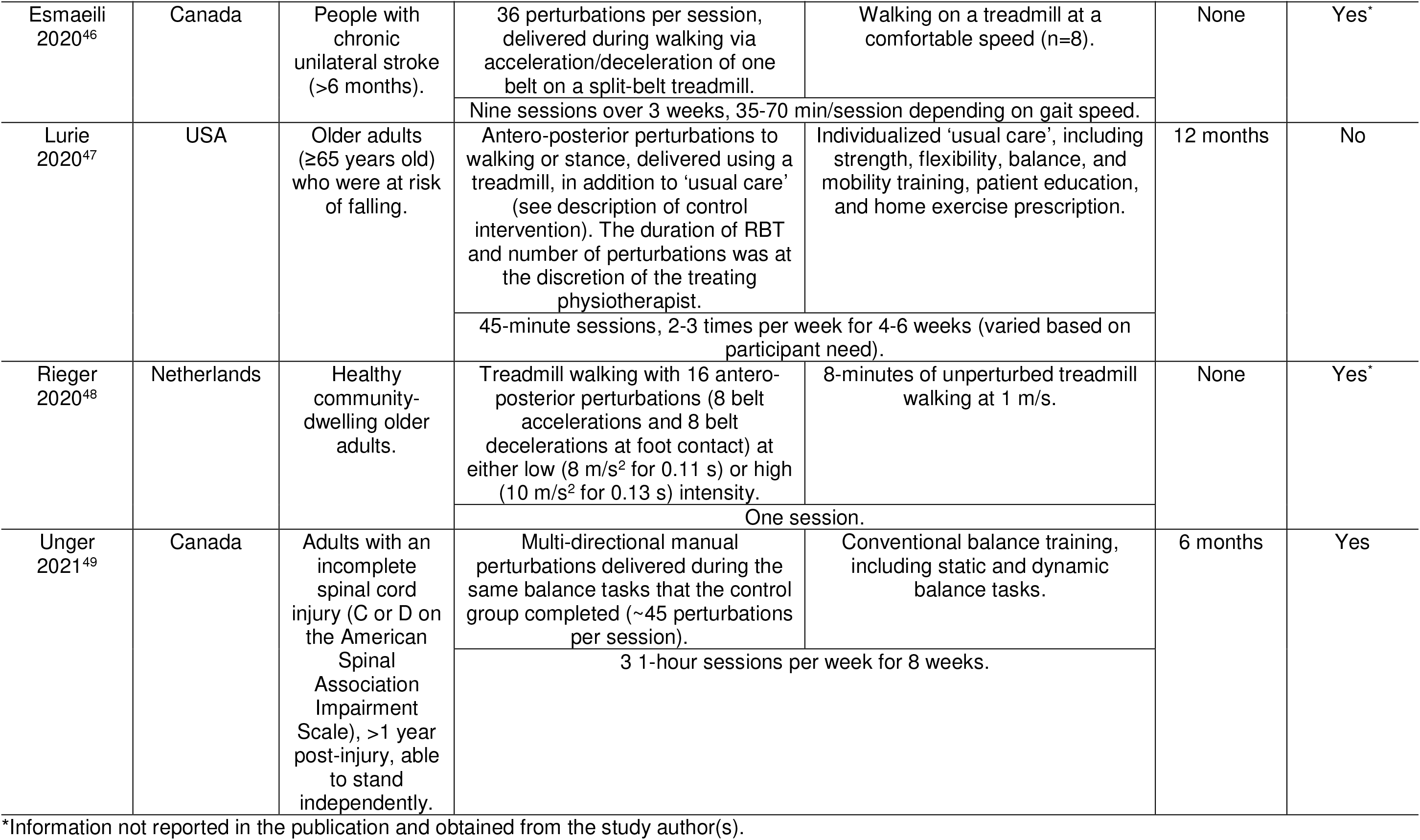
Characteristics of included studies

**Figure 1:**
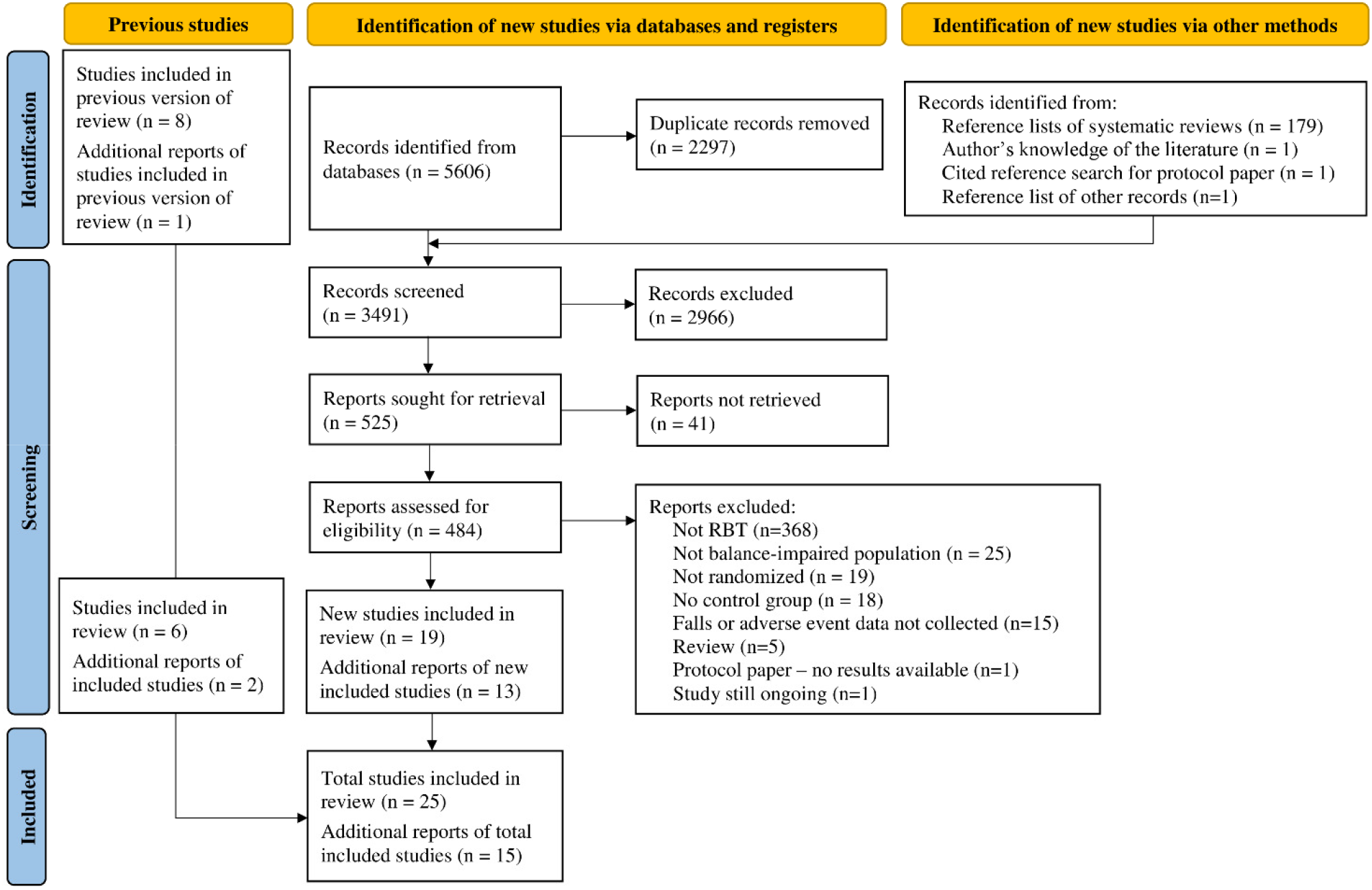
PRISMA flow diagram

### Study characteristics

Participants in studies included in this review were community-living older adults;^9, 31, 33-35, 40, 45, 47, 48, 51^ adults with Parkinson’s disease,^27, 29, 32, 37, 38, 41, 42^ stroke,^8, 30, 39, 44, 46^ cerebral palsy,^36^ and spinal cord injury;^49^ and long-term care residents.^28, 43^ All studies had RBT as either a sole or a significant component of the experimental intervention. Three studies did not provide any intervention to the control group,^27, 29, 34^ and the remaining studies used non-RBT exercise for the control intervention. The loss of balance during RBT was caused by manual perturbations,^8, 27, 30, 32, 34, 36, 37, 39, 41, 49^challenging balance tasks to evoke internal perturbations,^8, 49^ single-axis or multi-directional perturbations on a treadmill or a moveable platform,^9, 28, 29, 38, 40, 42-48^ and/or slip and trip perturbations on a walkway with movable tiles/platforms and tripping obstacles.^33, 35, 45^ The frequency of RBT ranged from one training session total^33, 35, 48^ to 3 times per week.^9, 27-32, 34, 41, 43, 47, 49^ The duration of RBT training ranged from 15 minutes^33^ to 1.5 hours^36^ per session; most studies (19/25) conducted RBT for 30 to 60 minutes per session.

### Methodological quality and risk of bias in studies

The methodological and reporting quality assessed using PEDro scale was good (≥ 6) in 18 studies, fair (4 to 5) in 6 studies, and poor (≤ 3) in 1 study (Table 2). Out of 25 studies, 19 reported conducting an intention-to-treat analysis. None of the included studies blinded participants to the intervention they received. In one study,^32^ the therapists did not know whether they were providing the experimental or control intervention.

**Table 2:** PEDro scores of included studies

| Study | PEDro items |  |  |  |  |  |  |  |  |  |  | Total |
| --- | --- | --- | --- | --- | --- | --- | --- | --- | --- | --- | --- | --- |
|  | 1* | 2 | 3 | 4 | 5 | 6 | 7 | 8 | 9 | 10 | 11 |  |
| Toole 2000<sup>27</sup> | No | 1 | 0 | 1 | 0 | 0 | 0 | 0 | 0 | 1 | 1 | 4 |
| Shimada 2004<sup>28</sup> | Yes | 1 | 0 | 1 | 0 | 0 | 1 | 0 | 1 | 1 | 1 | 6 |
| Marigold 2005<sup>30</sup> | Yes | 1 | 1 | 1 | 0 | 0 | 1 | 0 | 1 | 1 | 1 | 7 |
| Protas 2005<sup>29</sup> | Yes | 1 | 0 | 1 | 0 | 0 | 0 | 0 | 1 | 1 | 0 | 4 |
| Maki 2008<sup>31</sup> | Yes | 1 | 0 | 0 | 0 | 0 | 0 | 0 | 1 | 0 | 0 | 2 |
| Mansfield 2010<sup>9</sup> | Yes | 1 | 1 | 1 | 0 | 0 | 0 | 1 | 0 | 0 | 0 | 4 |
| Smania 2010<sup>32</sup> | Yes | 1 | 1 | 1 | 0 | 0 | 1 | 0 | 1 | 1 | 1 | 7 |
| Parijat 2012<sup>33</sup> | Yes | 1 | 0 | 1 | 0 | 0 | 0 | 0 | 0 | 1 | 1 | 4 |
| Halvarsson 2013<sup>34</sup> | Yes | 1 | 1 | 1 | 0 | 0 | 1 | 1 | 1 | 1 | 1 | 8 |
| Pai 2014<sup>35</sup> | Yes | 1 | 0 | 1 | 0 | 0 | 0 | 0 | 1 | 1 | 1 | 5 |
| Morgan 2015<sup>36</sup> | Yes | 1 | 1 | 1 | 0 | 0 | 1 | 1 | 1 | 1 | 1 | 8 |
| Schlenstedt 2015<sup>37</sup> | Yes | 1 | 0 | 1 | 0 | 0 | 1 | 0 | 1 | 1 | 1 | 6 |
| Shen 2015<sup>38</sup> | Yes | 1 | 1 | 1 | 0 | 0 | 1 | 0 | 1 | 1 | 1 | 7 |
| Kumar 2016<sup>39</sup> | Yes | 1 | 0 | 1 | 0 | 0 | 0 | 0 | 0 | 1 | 1 | 4 |
| Kurz 2016<sup>40</sup> | Yes | 1 | 1 | 0 | 0 | 0 | 1 | 0 | 1 | 1 | 1 | 6 |
| Gandolfi 2017<sup>41</sup> | Yes | 1 | 0 | 1 | 0 | 0 | 1 | 1 | 0 | 1 | 1 | 6 |
| Steib 2017<sup>42</sup> | Yes | 1 | 1 | 1 | 0 | 0 | 1 | 1 | 0 | 1 | 1 | 7 |
| Mansfield 2018<sup>8</sup> | Yes | 1 | 1 | 1 | 0 | 0 | 1 | 1 | 1 | 1 | 1 | 8 |
| Aviles 2019<sup>43</sup> | Yes | 1 | 0 | 1 | 0 | 0 | 1 | 0 | 1 | 1 | 1 | 6 |
| Handelzalts 2019<sup>44</sup> | Yes | 1 | 1 | 1 | 0 | 0 | 1 | 1 | 1 | 1 | 1 | 8 |
| Okubo 2019<sup>45</sup> | Yes | 1 | 1 | 1 | 0 | 0 | 1 | 1 | 1 | 1 | 1 | 8 |
| Esmaili 2020<sup>46</sup> | Yes | 1 | 0 | 1 | 0 | 0 | 1 | 1 | 1 | 1 | 1 | 7 |
| Lurie 2020<sup>47</sup> | Yes | 1 | 1 | 0 | 0 | 0 | 0 | 1 | 1 | 1 | 1 | 6 |
| Rieger 2020<sup>48</sup> | Yes | 1 | 0 | 1 | 0 | 0 | 0 | 1 | 1 | 1 | 1 | 6 |
| Unger 2021<sup>49</sup> | Yes | 1 | 1 | 1 | 0 | 0 | 1 | 1 | 1 | 1 | 1 | 8 |

PEDro items: (1) eligibility criteria; (2) random allocation; (3) concealed allocation; (4) baseline comparability; (5) blind subjects; (6) blind therapists; (7) blind assessors; (8) adequate follow-up; (9) intention-to-treat analysis; (10) between-group comparisons; (11) point estimates and variability. \*The eligibility criteria item in the PEDro scale does not contribute to the PEDro score. PEDro: Physiotherapy Evidence Database.

Risk of bias, assessed using the revised Cochrane Risk of Bias Tool^17, 18^ in the studies that reported risk of falls and rate of falls (n=14 in total), was high in 9 studies, found to have some concerns for risk of bias in 1 study, and low in 4 studies (Table 3). Considering the findings from each of the five domains, a high risk of bias arose from the randomization process (5/14), due to deviations from the intended interventions (2/14), missing outcome data (7/14), and due to a risk of bias in measurement of the outcome (5/14).

**Table 3:** Risk of bias assessments using the revised Cochrane Risk of Bias Tool in studies with information about risk of falls and rate of falls

| Study | Domain 1 | Domain 2a | Domain 2b | Domain 3 | Domain 4 | Domain 5 | Overall risk of bias |
| --- | --- | --- | --- | --- | --- | --- | --- |
| Aviles 2019<sup>43</sup> | High | Some concerns | Some concerns | High | Low | Low | High |
| Shimada 2004<sup>28</sup> | High | Low | Some concerns | High | Low | Low | High |
| Marigold 2005<sup>30</sup> | Low | Low | Some concerns | High | Low | Low | High |
| Protas 2005<sup>29</sup> | High | High | Low | Low | Some concerns | Low | High |
| Maki 2008<sup>31</sup> | High | Some concerns | High | High | High | Low | High |
| Mansfield 2010<sup>9</sup> | Low | Low | Low | Low | High | Low | High |
| Smania 2010<sup>32</sup> | Low | Low | Low | Low | Low | Low | Low |
| Halvarsson 2013<sup>34</sup> | Low | Low | Some concerns | High | Some concerns | Low | High |
| Pai 2014<sup>35</sup> | High | Low | Low | High | High | Low | High |
| Shen 2015<sup>38</sup> | Low | Low | Low | Low | Low | Low | Low |
| Kurz 2016<sup>40</sup> | Some concerns | Low | Low | Low | Low | Low | Some concerns |
| Mansfield 2018<sup>8</sup> | Low | Low | Low | Low | Low | Low | Low |
| Lurie 2020<sup>47</sup> | Low | Low | Low | Low | High | Low | High |
| Unger 2021<sup>49</sup> | Low | Low | Low | Low | Low | Low | Low |

D: domain; D1: Risk of bias arising from the randomization process; D2a: Risk of bias due to deviations from the intended interventions (effect of assignment to intervention); D2b: Risk of bias due to deviations from the intended interventions (effect of adhering to intervention); D3: Missing outcome data; D4: Risk of bias in measurement of the outcome; D5: Risk of bias in selection of the reported result.

## Results of synthesis

### Effect of RBT on falls in daily life

Fourteen studies monitored falls in daily life after the intervention (Table 1); the falls monitoring duration ranged from 2 weeks to 1 year. There were 1,049 participants in 13 studies that reported the number of participants who experienced 1 or more falls in daily life after the intervention (539 were assigned to the RBT groups and 510 to the control groups). Ten of the 13 studies with fall risk data reported that fewer participants in the RBT groups experienced falls following training compared to the control groups (Table 4). Participants who completed RBT were less likely to fall than those in the control group; the overall risk ratio for all 13 studies combined was 0.75 (95% CI=0.60, 0.92; p=0.006, I^2^=37%; Figure 2a). The risk ratio was unchanged when only studies with an active exercise control intervention were included in the analysis (11 studies; risk ratio=0.75; 95% CI=0.64, 0.88; p=0.0004, I^2^=11%; Figure 3).

**Table 4:**
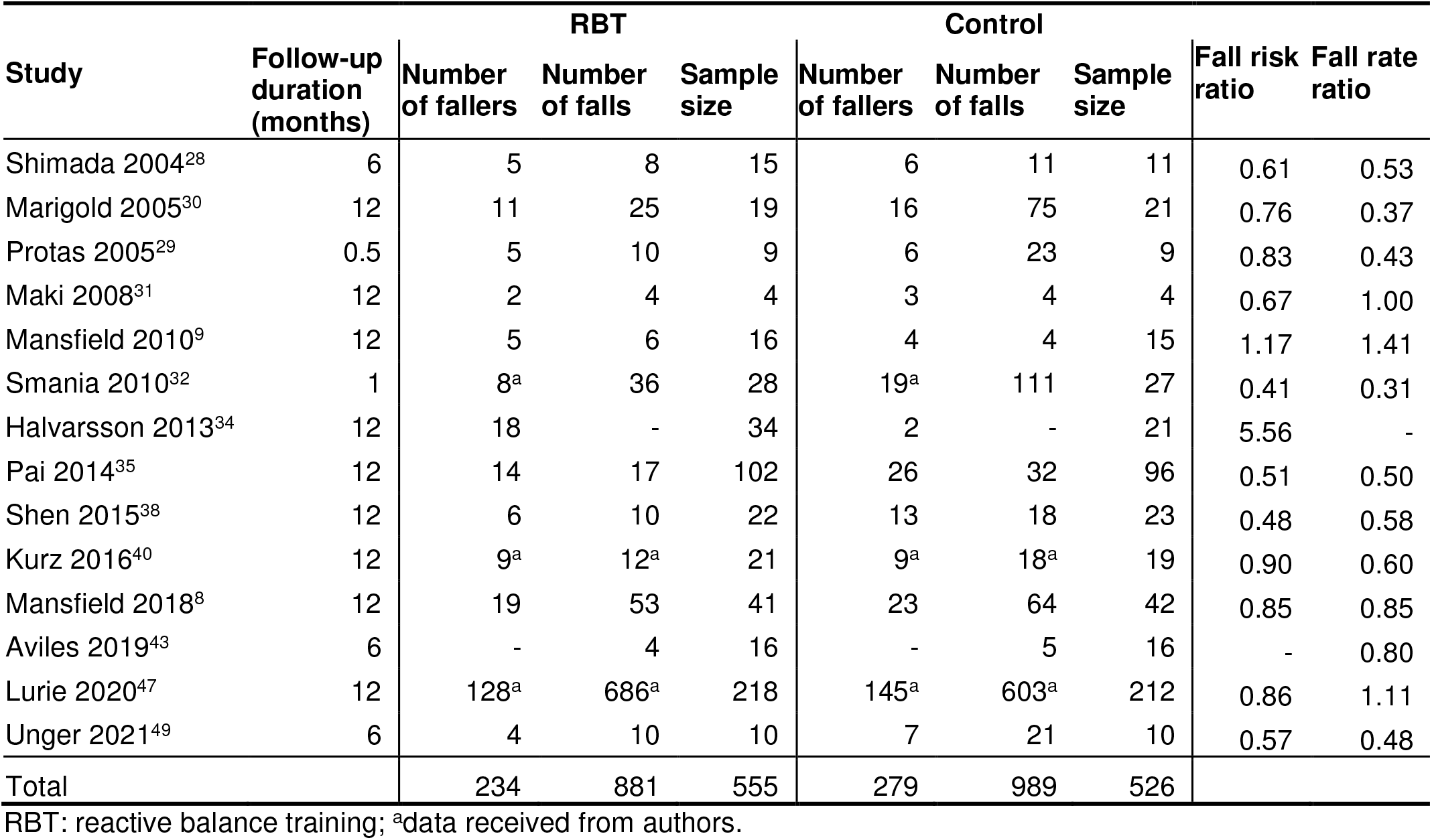
Falls after the intervention in the included studies

| Study | Follow-up duration (months) | RBT |  |  | Control |  |  | Fall risk ratio | Fall rate ratio |
| --- | --- | --- | --- | --- | --- | --- | --- | --- | --- |
|  |  | Number of fallers | Number of falls | Sample size | Number of fallers | Number of falls | Sample size |  |  |
| Shimada 2004<sup>28</sup> | 6 | 5 | 8 | 15 | 6 | 11 | 11 | 0.61 | 0.53 |
| Marigold 2005<sup>30</sup> | 12 | 11 | 25 | 19 | 16 | 75 | 21 | 0.76 | 0.37 |
| Protas 2005<sup>29</sup> | 0.5 | 5 | 10 | 9 | 6 | 23 | 9 | 0.83 | 0.43 |
| Maki 2008<sup>31</sup> | 12 | 2 | 4 | 4 | 3 | 4 | 4 | 0.67 | 1.00 |
| Mansfield 2010<sup>9</sup> | 12 | 5 | 6 | 16 | 4 | 4 | 15 | 1.17 | 1.41 |
| Smania 2010<sup>32</sup> | 1 | 8<sup>a</sup> | 36 | 28 | 19<sup>a</sup> | 111 | 27 | 0.41 | 0.31 |
| Halvarsson 2013<sup>34</sup> | 12 | 18 | - | 34 | 2 | - | 21 | 5.56 | - |
| Pai 2014<sup>35</sup> | 12 | 14 | 17 | 102 | 26 | 32 | 96 | 0.51 | 0.50 |
| Shen 2015<sup>38</sup> | 12 | 6 | 10 | 22 | 13 | 18 | 23 | 0.48 | 0.58 |
| Kurz 2016<sup>40</sup> | 12 | 9<sup>a</sup> | 12<sup>a</sup> | 21 | 9<sup>a</sup> | 18<sup>a</sup> | 19 | 0.90 | 0.60 |
| Mansfield 2018<sup>8</sup> | 12 | 19 | 53 | 41 | 23 | 64 | 42 | 0.85 | 0.85 |
| Aviles 2019<sup>43</sup> | 6 | - | 4 | 16 | - | 5 | 16 | - | 0.80 |
| Lurie 2020<sup>47</sup> | 12 | 128<sup>a</sup> | 686<sup>a</sup> | 218 | 145<sup>a</sup> | 603<sup>a</sup> | 212 | 0.86 | 1.11 |
| Unger 2021<sup>49</sup> | 6 | 4 | 10 | 10 | 7 | 21 | 10 | 0.57 | 0.48 |
| Total |  | 234 | 881 | 555 | 279 | 989 | 526 |  |  |

RBT: reactive balance training; <sup>a</sup>data received from authors. **Figure 2:** Results of meta-analysis for risk of falls (Panel A) and rate of falls (Panel B).

**Figure 2:**
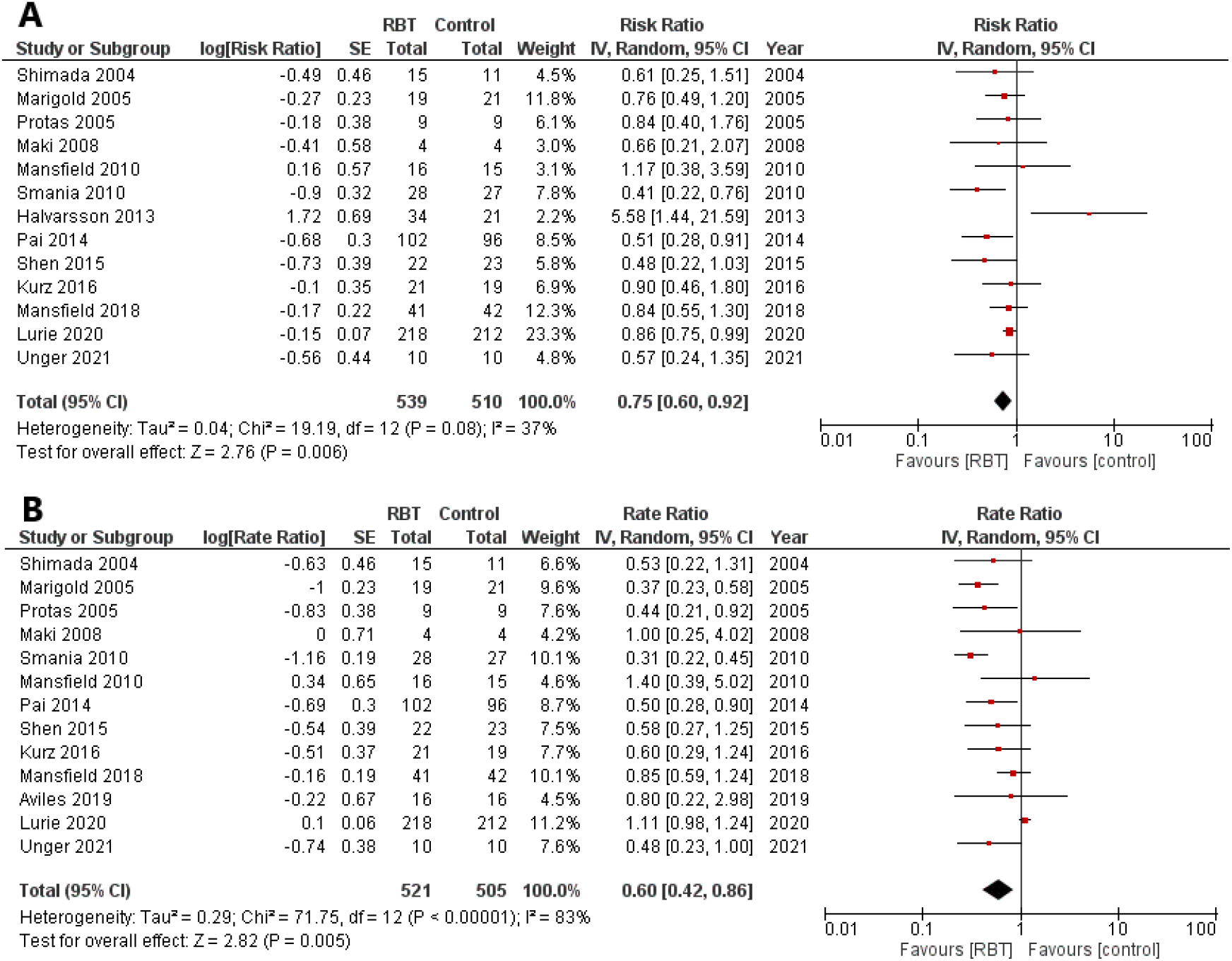
Results of meta-analysis for risk of falls (Panel A) and rate of falls (Panel B).

**Figure 3:**
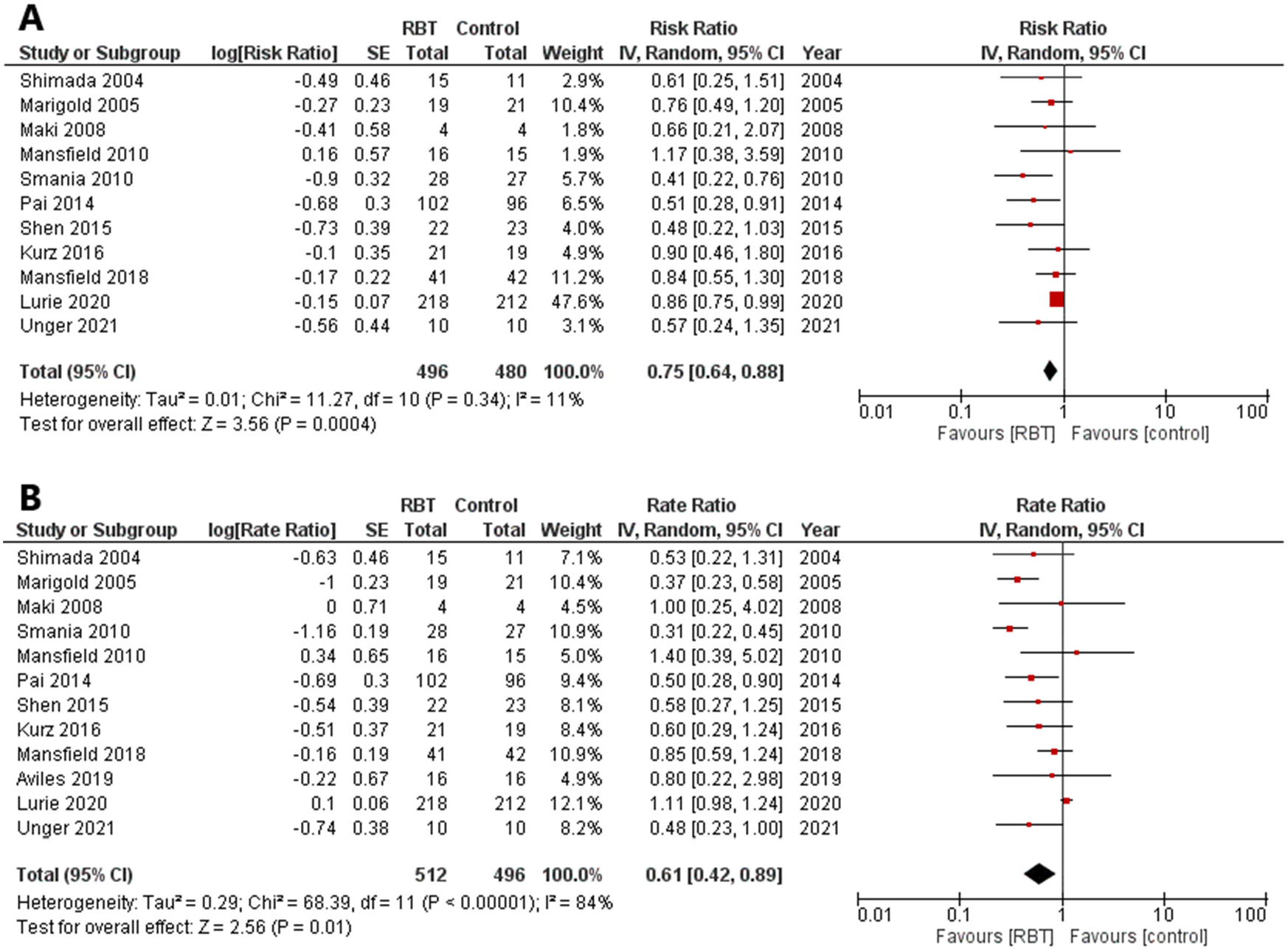
Results of meta-analysis for risk of falls (Panel A) and rate of falls (Panel B) in studies with an active non-RBT control intervention

There were 1,026 participants in 13 studies that reported the number of falls experienced by participants in daily life after completing the intervention (521 were assigned to RBT groups, and 505 to the control groups). Ten of the 13 studies with fall rate data reported fewer falls in the RBT groups compared to the control groups (Table 4); the overall rate ratio for all 13 studies combined was 0.60 (95% CI=0.42, 0.86; p=0.005, I^2^=83%; Figure 2b). Again, the rate ratio was unchanged when only studies with an active exercise control intervention were included in the analysis (12 studies; rate ratio=0.61; 95% CI=0.42, 0.89; p=0.01, I^2^=84%; Figure 3).

### Adverse events

Nineteen studies monitored adverse events. The nature, frequency, and severity of adverse events reported during RBT and control interventions are presented in Table 5. During RBT, participants reported pain (14.4%), subjective or psychological events (5.2%), musculoskeletal events (4.4%), neurological events (2.6%), fatigue (1.8%), and other adverse events (0.7%) such as fall with no injury during training or illness during training (Table 5). During control interventions, participants reported pain (9.9%), musculoskeletal events (4.6%), subjective or psychological events (1.9%), fatigue (1.1%), cardiorespiratory events (1.1%), and neurological events (0.8%; Table 5). Among 271 RBT participants, 79 reported adverse events (mild: n=63, moderate: n=15, severe: n=1; Table 5). Among 263 control participants, 51 reported adverse events (mild: n=43, moderate: n=8). There were more participants reporting adverse events in RBT groups compared to control groups (RBT: 29%, control: 19%; p=0.018).

**Table 5:** Number of participants with adverse events in each intervention group

| Category | Nature of adverse events | RBT (n=271) |  |  |  | Control (n=263) |  |  |  |
| --- | --- | --- | --- | --- | --- | --- | --- | --- | --- |
|  |  | Mild | Moderate | Severe | Total | Mild | Moderate | Severe | Total |
| Cardio-respiratory | Shortness of breath | 0 | 0 | 0 | 0 | 2 | 0 | 0 | 2 |
|  | Abnormal cardiovascular response to exercise | 0 | 0 | 0 | 0 | 0 | 1 | 0 | 1 |
| Fatigue |  | 5 | 0 | 0 | 5 | 3 | 0 | 0 | 3 |
| Musculo-skeletal | Muscle fatigue / soreness | 6 | 0 | 0 | 6 | 12 | 0 | 0 | 12 |
|  | Injury during training | 3 | 2 | 1 | 6 | 0 | 0 | 0 | 0 |
| Neurological | Dizziness | 5 | 0 | 0 | 5 | 2 | 0 | 0 | 2 |
|  | Seizure | 1 | 0 | 0 | 1 | 0 | 0 | 0 | 0 |
|  | Headache | 1 | 0 | 0 | 1 | 0 | 0 | 0 | 0 |
| Pain | Non-specified pain | 20 | 0 | 0 | 20 | 19 | 0 | 0 | 19 |
|  | Upper extremity pain | 1 | 0 | 0 | 1 | 0 | 0 | 0 | 0 |
|  | Lower extremity pain | 13 | 3 | 0 | 16 | 4 | 3 | 0 | 7 |
|  | Back pain | 1 | 1 | 0 | 2 | 0 | 0 | 0 | 0 |
| Subjective or psychological | Anxiety during training | 5 | 1 | 0 | 6 | 0 | 0 | 0 | 0 |
|  | Did not like intervention | 0 | 6 | 0 | 6 | 1 | 4 | 0 | 5 |
|  | Falls due to increased risk taking | 1 | 0 | 0 | 1 | 0 | 0 | 0 | 0 |
|  | Fear of injury | 0 | 1 | 0 | 1 | 0 | 0 | 0 | 0 |
| Other | Fall during training with no injury | 1 | 0 | 0 | 1 | 0 | 0 | 0 | 0 |
|  | Sick during training period | 0 | 1 | 0 | 1 | 0 | 0 | 0 | 0 |
| Total |  | 63 | 15 | 1 | 79 | 43 | 8 | 0 | 51 |

RBT: reactive balance training; n: total number of participants in the respective study groups that reported adverse events.

### Reporting bias

Funnels plots^21^ were examined as per the Sterne *et al*.^20^ recommendations to assess reporting bias in the results of meta-analysis of fall risk and rate (Figure 4) The asymmetry in the funnel plots was likely due to a possibility of selective outcome reporting,^28, 29^ the true clinical heterogeneity of studies,^9, 28, 29, 32, 43^ and a random chance.^34^

**Figure 4:**
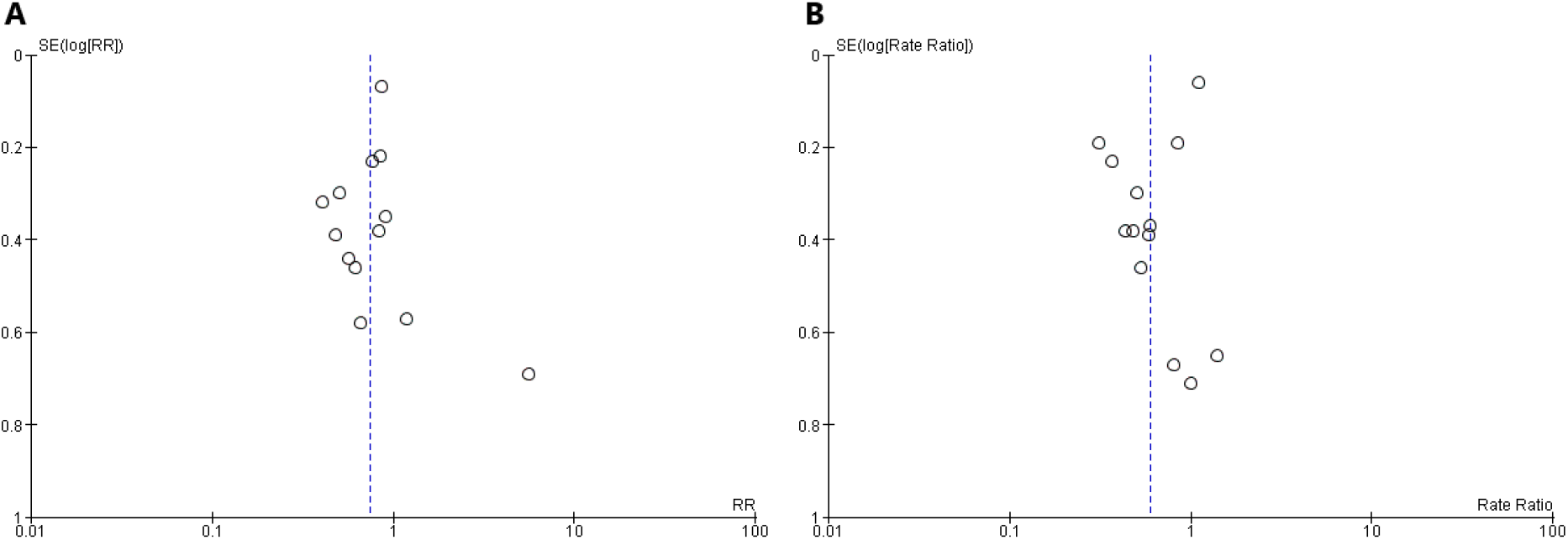
Funnel plot of meta-analysis for risk of falls (Panel A) and rate of falls (Panel B).

### Certainty of evidence

GRADE certainty of evidence was low to moderate for risk of falls and rate of falls (Table 6), due to a lack of concealed allocation, a lack of blinding procedures, possibility of selective outcome reporting, and an incomplete outcome data due to attrition in the included studies.

**Table 6:** Certainty assessment using GRADE recommendations

| Certainty assessment |  |  |  |  |  |  | Effect | Certainty |
| --- | --- | --- | --- | --- | --- | --- | --- | --- |
| Number of studies | Study design | Risk of bias | Inconsistency | Indirectness | Imprecision | Other considerations | Relative (95% CI) |  |
| **Risk ratio** |  |  |  |  |  |  |  |  |
| 13 | randomised trials | serious <sup>a</sup> | serious <sup>c</sup> | not serious | not serious | none | **Risk ratio 0.75**<br>(0.60 to 0.92) | ⊕⊕○○<br>LOW |
| **Risk ratio with active control** |  |  |  |  |  |  |  |  |
| 11 | randomised trials | serious <sup>b</sup> | not serious | not serious | not serious | none | **Risk ratio 0.75**<br>(0.64 to 0.88) | ⊕⊕⊕○<br>MODERATE |
| **Rate ratio** |  |  |  |  |  |  |  |  |
| 13 | randomised trials | serious <sup>a</sup> | serious <sup>c</sup> | not serious | not serious | none | **Rate ratio 0.60**<br>(0.42 to 0.86) | ⊕⊕○○<br>LOW |
| **Rate ratio with active control** |  |  |  |  |  |  |  |  |
| 12 | randomised trials | serious <sup>b</sup> | serious <sup>c</sup> | not serious | not serious | none | **Rate ratio 0.61**<br>(0.42 to 0.89) | ⊕⊕○○<br>LOW |

RBT: Reactive balance training; CI: Confidence interval; RR: Risk ratio; a: Shimada et al<sup>28</sup> - No concealed allocation, No blinding of participants and therapists, No intention to treat analysis, Possibility of selective reporting; Protas et al<sup>29</sup> - No concealed allocation, No blinding of participants, No intention to treat analysis, Possibility of selective reporting; Pai et al<sup>35</sup> - No concealed allocation, No blinding of participants and therapists, No intention to treat analysis; b: Shimada et al<sup>28</sup> - No concealed allocation, No blinding of participants and therapists, No intention to treat analysis, Possibility of selective reporting; c: I<sup>2</sup>>30%.

## DISCUSSION

This study aimed to determine the effect of RBT on falls in daily life. Overall, we found moderate certainty of evidence that RBT can reduce falls in daily life among individuals at increased risk of falls. The overall effect for fall risk (ratio = 0.75) indicates that those who completed RBT had a 25% reduction in the risk of experiencing one or more falls compared to controls. The overall effect for fall rate (ratio = 0.60) indicates that those who completed RBT experienced 40% fewer falls when compared to controls. These risk and rate ratios are similar to our previous meta-analysis of RBT on falls in daily life, which only included 8 studies (risk ratio: 0.71, rate ratio: 0.54).^7^ Likewise, in a meta-analysis of 7 studies, Okubo et al^53^ reported that reactive stepping interventions reduced falls among older adults with increased risk of falls (risk ratio: 0.60, rate ratio: 0.52). Repeated task-specific motor experience, such as reactive stepping, is necessary to learn perceptual-motor abilities that are essential for recovery from loss of balance.^54^ Since avoiding a fall requires the whole body to react, exercise interventions that intend to reduce the risk of falls must evoke task-specific total body balance reactions. A large Cochrane review^3^ including 25,160 individuals aged 60+ years from 116 studies reported that interventions with a total weekly dose of 3+ hours of task-specific balance and functional exercises were effective in reducing falls (rate ratio: 0.58) compared to controls. These findings suggest that task-specific tailored RBT programs with well-defined dosage prescription should be developed for people with balance impairments.

Unlike other types of exercise, dosage is not well defined in balance prescription. Specifically, the intensity of balance training is often assessed using metrics that are not appropriate for balance training, such as the participant’s rate of perceived exertion and/or the therapist’s perception of the participant’s ability (i.e., safety).^55^ For example, Halvarsson *et al*.^34^ prescribed exercises at five different levels with each reflecting progressive demands on the postural control system, whereas Smania *et al*.^32^ challenged participants during each session with 10 different exercises before increasing the complexity of the tasks; however, both studies did not quantify the perturbation intensity. It is possible that the dosage of RBT was not sufficient in some studies included in our review to impact fall risk in daily life.^56^ Furthermore, the amount of training has to reach an asymptotic level within one session in order to demonstrate performance gains across sessions.^57^ Therefore, there is a need to characterize the intensity of RBT (such as perturbation force) and determine dose-response effect of RBT in future studies at different stages of recovery from balance impairments.

Although the methodological and reporting quality of the included studies was good on average, there was a high risk of bias arising from the randomization process, missing outcome data, and bias in measurement of the outcome. Not blinding therapists who deliver interventions could introduce bias in the treatment effects.^58^ However, it is not possible to blind therapists in exercise studies where the therapist delivers the exercise intervention. One study attempted to blind the therapists to aims of the study,^32^ but the therapists were still aware of the nature and specific details of the intervention. Furthermore, having different therapists provide the two interventions, as was the case for Smania *et al*.,^32^ may introduce another source of variability between groups (i.e., the therapists may differ in their treatment approaches).

The prevalence of adverse events was higher during RBT compared to control interventions. The between-group difference in adverse events seemed to be greatest for subjective or psychological events, although these data were not statistically analysed due to the low numbers of each type of adverse event. Seven RBT participants reported fear or anxiety related to the perturbations, whereas no control participants reported fear or anxiety related to the control intervention. This finding highlights the need to build trust between the therapist and client to alleviate these anxieties.^59^ Therapists report that, as clients gradually overcome these anxieties with training, RBT can help to build self-efficacy and a sense of achievement.^59^

Findings regarding adverse events should be interpreted with caution. Fifteen studies that met most of our inclusion criteria were not included in the review because they did not monitor adverse events, and six trials that were included in the current review did not monitor adverse events. Therefore, approximately half of RBT trials do not monitor or report adverse events. This is consistent with two other reviews, where only 43 and 44% of exercise interventions reported adverse events.^60, 61^ Even among those trials that did report adverse events, it is possible that not all events were reported; some study authors only report certain types of adverse events or only report adverse events above a certain threshold (e.g., only severe adverse events, or only those events reported by a certain proportion of participants).^62-65^ Seven RBT studies that monitored adverse events did not report any; given the known adverse events associated with exercise, it is likely that low or reports of zero adverse events in exercise studies are either due to selective non-reporting or under-reporting of adverse events.

Control interventions may have been less ‘intense’ than RBT. For example, Mansfield *et al*.^9^ and Marigold *et al*.^30^ used low intensity activities like stretching or relaxation for the control groups in their studies. In Okubo *et al*.’s^53^ study, RBT participants completed 120 minutes of training on a trip and slip walkway with 30 trips and slips in 3 sessions, while control participants completed 40 minutes of walking on the same path with no perturbations in 1 session. Likewise, Shimada *et al*.^28^ and Lurie *et al*.^47^ assigned ‘usual care’ to the control groups while conducting RBT in addition to ‘usual care’ for the experimental groups. Since the control interventions were often less challenging than RBT, with either lower dose of therapy or no intervention, the effect size estimate for RBT on fall risk could be inflated. Furthermore, the lower intensity or duration of control interventions compared to RBT could, in part, explain the increased prevalence of adverse events during RBT compared to control interventions. Future RBT studies should take steps to include clinically meaningful control interventions that are comparable to the experimental intervention while investigating the effect of RBT on fall risk.^66^ In addition, it is necessary to follow best practices for adverse event reporting to learn about both the risks and benefits of RBT for people with balance impairments.^67^

## Limitations

We noted asymmetry in the funnel plots generated to determine the presence of biases in meta-analysis of fall risk and rate. However, we were not able to confirm whether the asymmetry in the funnel plots was due to publication bias. Funnel plots determine whether studies with lower precision (e.g., higher standard error) differ from studies with higher precision (lower standard error).^68^ The standard error was the highest in the study by Halvarsson *et al*.^34^ compared to the 12 other studies included in the meta-analysis of risk of falls. In this study,^34^ participants performed ‘typical’ balance exercises with a focus on maintaining balance, and also experienced external perturbations; which were provided by the instructor or partner. The authors did not report the prescribed intensity of the perturbations.^34^ This could have been a major factor (clinical heterogeneity) influencing the effect of RBT on the risk of falls in daily life.^69^ Because of the above-mentioned factor and since none of the tests available for publication bias have been compared against a gold standard,^70^ we were not able to assess publication bias in our study. Therefore, our confidence in the estimate of the reduction in the risk and rate of falls with RBT is ‘low’ to support a recommendation. Further research that addresses the limitations highlighted in this study is likely to change the estimate.

We were unable to confirm the prevalence of adverse events for six studies. From our correspondence with study authors, we noted that several studies did not report mild and/or expected adverse events associated with exercise in the publication. It is possible that most of the minor intervention-related adverse events, such as muscle or joint pain, were reported in the following session (i.e., the participants are asked “how did you feel after the last session?”); therefore, there is a lack of certainty about adverse events for studies with only one session or without longer term intervention. Such under-reporting of adverse events could have biased our results.

## Conclusions

Older adults and individuals with balance impairments had a lower likelihood of falls in daily life after participating in RBT compared to control interventions. The certainty of evidence was ‘low’ to ‘moderate’ due to a lack of concealed allocation, a lack of blinding procedures, a possibility of selective outcome reporting, and an incomplete outcome data due to attrition, indicating the need for studies with a lower risk of bias. Participants of RBT reported more adverse events than control groups; however, the prevalence of adverse events for both RBT and control groups is likely underestimated as only about half of RBT studies monitored adverse events.

## Data Availability

All data produced in the present work are contained in the manuscript.

## Acknowledgements

We thank Jessica Babineau, BA, MLIS, for developing the search strategy and conducting the literature search. Avril Mansfield was supported by a New Investigator Award from the Canadian Institutes of Health Research.

## Appendix A: Sample search strategy (Ovid MEDLINE)

1. (perturb* adj3 (train* or rehab* or exercis*)).tw,kf. (354)
2. (platform* adj2 (train* or rehab* or exercis*)).tw,kf. (554)
3. (surface translation? adj2 (train* or rehab* or exercis*)).tw,kf. (1)
4. (“dynamic balanc*” adj4 (train* or rehab* or exercis*)).tw,kf. (203)
5. (“dynamic stabil*” adj4 (train* or rehab* or exercis*)).tw,kf. (45)
6. (“reactive balanc*” adj4 (train* or rehab* or exercis*)).tw,kf. (32)
7. ((slip? or slipping) adj2 (train* or rehab* or exercis*)).tw,kf. (45)
8. ((trip? or tripping) adj2 (train* or rehab* or exercis*)).tw,kf. (53)
9. 9 1 or 2 or 3 or 4 or 5 or 6 or 7 or 8 (1222)
10. ((step? or stepping) adj4 (train* or rehab* or exercis*)).tw,kf. (3211)
11. (gait adj4 (train* or rehab* or exercis*)).tw,kf. (3529)
12. ((walk? or walking) adj4 (train* or rehab* or exercis*)).tw,kf. (5868)
13. (locomot* adj4 (train* or rehab* or exercis*)).tw,kf. (1271)
14. (balanc* adj4 (train* or rehab* or exercis*)).tw,kf. (5701)
15. (stabil* adj4 (train* or rehab* or exercis*)).tw,kf. (2422)
16. (agil* adj4 (train* or rehab* or exercis*)).tw,kf. (260)
17. 10 or 11 or 12 or 13 or 14 or 15 or 16 (20144)
18. perturb*.tw,kf. (111319)
19. platform*.tw,kf. (167742)
20. surface translation?.tw,kf. (163)
21. destabil*.tw,kf. (34912)
22. compensat*.tw,kf. (160601)
23. react*.tw,kf. (1835834)
24. dynamic balanc*.tw,kf. (3340)
25. dynamic stabil*.tw,kf. (2075)
26. 18 or 19 or 20 or 21 or 22 or 23 or 24 or 25 (2266570)
27. 17 and 26 (2676)
28. 9 or 27 (3490)
29. randomized controlled trial.pt. (516491)
30. controlled clinical trial.pt. (93915)
31. random*.ab. (1136957)
32. placebo.ab. (212267)
33. trial.ab. (526103)
34. groups.ab. (2111952)
35. 29 or 30 or 31 or 32 or 33 or 34 (3271056)
36. exp animals/ not humans.sh. (4753439)
37. 35 not 36 (2799327)
38. 28 and 37 (1498)
39. limit 38 to english language (1464)

## Appendix B List of all records used for included studies

***Toole 2000*** (Trial registration: not reported)

- Toole T, Hirsch MA, Forkink A, Lehman DA, Maitland CG. The effects of a balance and strength training program on equilibrium in Parkinsonism: a preliminary study. *Neurorehabilitation*. 2000;14(3):165-174. (main publication)

***Shimada 2004*** (Trial registration: not reported)

- Shimada H, Obuchi S, Furuna T, Suzuki T. New intervention program for preventing falls among frail elderly people: the effects of perturbed walking exercise using a bilateral separated treadmill. *Am J Phys Med Rehabil*. 2004;83(7):493-499. (main publication)

***Marigold 2005*** (Trial registration: Not reported

- Marigold DS, Eng JJ, Dawson AS, Inglis JT, Harris JE, Gylfadottir S. Exercise leads to faster postural reflexes, improved balance and mobility, and fewer falls in older persons with chronic stroke. *J Am Geriatr Soc*. 2005;53(3):416-423. (main publication)

***Protas 2005*** (Trial registration: not reported)

- Protas EJ, Mitchell K, Williams A, Qureshy H, Caroline K, Lai EC. Gait and step training to reduce falls in Parkinson’s disease. *Neurorehabilitation*. 2005;20(3):183-190. (main publication)

***Maki 2008*** (Trial registration: not reported)

- Maki BE, Cheng KCC, Mansfield A, et al. Preventing falls in older adults: new interventions to promote more effective change-in-support balance reactions. *J Electromyogr Kinesiol*. 2008;18(2):243-254. (main publication)

***Mansfield 2010*** (Trial registration: NCT00187317)

- Mansfield A, Peters AL, Liu BA, Maki BE. Effect of a perturbation-based balance training program on compensatory stepping and grasping reactions in older adults: a randomized controlled trial. *Phys Ther*. 2010;90(4):476-491. (main publication)
- Mansfield A, Peters AL, Liu BA, Maki BE. A perturbation-based balance training program for older adults: study protocol for a randomised controlled trial. *BMC Geriatr*. 2007;7:12. (protocol)
- Mansfield A. Development and evaluation of a perturbation-based balance-training program for older adults. Dissertation/ thesis. 2007;216. (secondary publication)

***Smania 2010*** (Trial registration: not reported)

- Smania N, Corato E, Tinazzi M, et al. Effect of balance training on postural instability in patients with idiopathic Parkinson’s disease. *Neurorehabil Neural Repair*. 2010;24(9):826-834. (main publication)

***Parijat 2012*** (Trial registration: not reported)

- Parijat P, Lockhart TE. Effects of moveable platform training in preventing slip-induced falls in older adults. *Ann Biomed Eng*. 2012;40(5):1111-1121. (main publication)

***Halvarsson 2013*** (Trial registration: not reported)

- Halvarsson A, Franzén E, Farén E, Olsson E, Oddsson L, Ståhle A. Long-term effects of new progressive group balance training for elderly people with increased risk of falling - a randomized controlled trial. *Clin Rehabil*. 2013;27(5):450-458. (main publication)
- Halvarsson A, Oddsson L, Olsson E, Faren E, Pettersson A, Stahle A. Effects of new, individually adjusted, progressive balance group training for elderly people with fear of falling and tend to fall: a randomized controlled trial. *Clin Rehabil*. 2011;25(11):1021-1031. (secondary publication)

***Pai 2014*** (Trial registration: not reported)

- Pai YC, Bhatt T, Yang F, Wang E. Perturbation training can reduce community-dwelling older adults’ annual fall risk: a randomized controlled trial. *J Gerontol A Biol Sci Med Sci*. 2014;69(12):1586-1594. (main publication)

***Morgan 2015*** (Trial registration: ACTRN12613000166774)

- Morgan P, Murphy A, Opheim A. The safety and feasibility of an intervention to improve balance dysfunction in ambulant adults with cerebral palsy: a pilot randomized controlled trial. *Clin Rehabil*. 2015;29(9):907-919. (main publication)

***Schlenstedt 2015*** (Trial registration: NCT02253563)

- Schlenstedt C, Paschen S, Kruse A, Raethjen J, Weisser B, Deuschl G. Resistance versus balance training to improve postural control in Parkinson’s disease: a randomized rater blinded controlled study. *PLoS ONE*. 2015;10(10):e0140584. (main publication)

***Shen 2015*** (Trial registration: not reported)

- Shen X, Mak MKY. Technology-assisted balance and gait training reduces falls in patients with Parkinson’s disease: a randomized controlled trial with 12-month follow-up. *Neurorehabil Neural Repair*. 2015;29(2):103-111. (main publication)
- Shen X, Mak MKY. Balance and gait training with augmented feedback improves balance confidence in people with Parkinson’s disease: a randomized controlled trial. *Neurorehabil Neural Repair*. 2014;28(6):524-535. (secondary publication)

***Kumar 2016*** (Trial registration: not reported)

- Kumar C, Pathan N. Effectiveness of manual perturbation exercises in improving balance, function and mobility in stroke patients: a randomized controlled trial. *J Nov Physiother*. 2016;6:2 (main publication)

***Kurz 2016*** (Trial registration: NCT01439451)

- Kurz I, Gimmon Y, Shapiro A, Debi R, Snir Y, Melzer I. Unexpected perturbations training improves balance control and voluntary stepping times in older adults - a double blind randomized control trial. *BMC Geriatr*. 2016;16:58. (main publication)
- Gimmon Y, Riemer R, Kurz I, Shapiro A, Debbi R, Melzer I. Perturbation exercises during treadmill walking improve pelvic and trunk motion in older adults: a randomized control trial. *Arch Gerontol Geriatr*. 2018;75:132-138. (secondary publication)

***Gandolfi 2017*** (Trial registration: not reported)

- Gandolfi M, Geroin C, Dimitrova E, et al. Virtual reality telerehabilitation for postural instability in Parkinson’s disease: a multicenter, single-blind, randomized, controlled trial. *BioMed Research International*. 2017(7962826) (main publication)

***Steib 2017*** (Trial registration: NCT01856244)

- Steib S, Klamroth S, Gassner H, et al. Perturbation during treadmill training improves dynamic balance and gait in Parkinson’s disease: a single-blind randomized controlled pilot trial. *Neurorehabil Neural Repair*. 2017;31(8):758-768. (main publication)
- Pasluosta CF, Steib S, Klamroth S, et al. Acute Neuromuscular adaptations in the postural control of patients with Parkinson’s disease after perturbed walking. *Front Aging Neurosci*. 2017;9:316. (secondary publication)
- Klamroth S, Gasner H, Winkler J, et al. Interindividual balance adaptations in response to perturbation treadmill training in persons with Parkinson disease. *J Neurol Phys Ther*. 2019;43(4):224-232. (secondary publication)
- Steib S, Klamroth S, Gasner H, et al. Exploring gait adaptations to perturbed and conventional treadmill training in Parkinson’s disease: time-course, sustainability, and transfer. *Hum Mov Sci*. 2019;64:123-132. (secondary publication)
- Klamroth S, Steib S, Gasner H, et al. Immediate effects of perturbation treadmill training on gait and postural control in patients with Parkinson’s disease. *Gait Posture*. 2016;50:102-108. (secondary publication)
- Gassner H, Steib S, Klamroth S, et al. Perturbation treadmill training improves clinical rating of the motor symptoms gait and postural stability, and sensor-based gait parameters in parkinson’s disease. *Gait Posture*. 2017;57:345-346. (secondary publication)
- Gassner H, Steib S, Klamroth S, et al. Perturbation treadmill training improves clinical characteristics of gait and balance in Parkinson’s disease. *J Parkinsons Dis*. 2019;9(2):413-426. (secondary publication)

***Mansfield 2018*** (Trial registration: ISRCTN05434601)

- Mansfield A, Aqui A, Danells CJ, et al. Does perturbation-based balance training prevent falls among individuals with chronic stroke? A randomised controlled trial. *BMJ Open*. 2018;8(8):e021510. (main publication)
- Mansfield A, Aqui A, Centen A, et al. Perturbation training to promote safe independent mobility post-stroke: study protocol for a randomized controlled trial. *BMC Neurol*. 2015;15:87. (protocol)
- Schinkel-Ivy A, Huntley AH, Aqui A, Mansfield A. Does perturbation-based balance training improve control of reactive stepping in individuals with chronic stroke? *J Stroke Cerebrovasc Dis*. 2019;28(4):935-943. (secondary publication)

***Aviles 2019*** (Trial registration: Not reported)

- Aviles J, Allin LJ, Alexander NB, Van Mullekom J, Nussbaum MA, Madigan ML. Comparison of treadmill trip-like training versus Tai Chi to improve reactive balance among independent older adult residents of senior housing: a pilot controlled trial. *J Gerontol A Biol Sci Med Sci*. 2019;74(9):1497-1503. (main publication)

***Handelzalts 2019*** (Trial registration: NCT02619175)

- Handelzalts S, Kenner-Furman M, Gray G, Soroker N, Shani G, Melzer I. Effects of perturbation-based balance training in subacute persons with stroke: a randomized controlled trial. *Neurorehabil Neural Repair*. 2019;33(3):213-224. (main publication)

***Okubo 2019*** (Trial registration: ACTRN12617000564358)

- Okubo Y, Sturnieks DL, Brodie MA, Duran L, Lord SR. Effect of reactive balance training involving repeated slips and trips on balance recovery among older adults: a blinded randomized controlled trial. *J Gerontol A Biol Sci Med Sci*. 2019;74(9):1489-1496. (main publication)

***Esmaeili 2020*** (Trial registration: NCT04314830)

- Esmaeili V, Juneau A, Dyer JO, et al. Intense and unpredictable perturbations during gait training improve dynamic balance abilities in chronic hemiparetic individuals: a randomized controlled pilot trial. *J Neuroeng Rehabil*. 2020;17(1):79. (main publication)

***Lurie 2020*** (Trial registration: NCT01006967)

- Lurie JD, Zagaria AB, Ellis L, et al. Surface perturbation training to prevent falls in older adults: a highly pragmatic, randomized controlled trial. *Phys Ther*. 2020;100(7):1153-1162. (main publication)
- Lurie JD, Zagaria AB, Pidgeon DM, Forman JL, Spratt KF. Pilot comparative effectiveness study of surface perturbation treadmill training to prevent falls in older adults. *BMC Geriatr*. 2013;13:49. (internal pilot study)

***Rieger 2020*** (Trial registration: not reported)

- Rieger MM, Papegaaij S, Pijnappels M, Steenbrink F, van Dieen JH. Transfer and retention effects of gait training with anterior-posterior perturbations to postural responses after medio-lateral gait perturbations in older adults. *Clin Biomech*. 2020;75:104988 (main publication)

***Unger 2021*** (Trial registration: NCT02960178)

- Unger J, Chan K, Lee JW, et al. The effect of perturbation-based balance training and conventional intensive balance training on reactive stepping ability in individuals with incomplete spinal cord injury or disease: a randomized clinical trial. *Front Neurol*. 2021;12:620367. (main publication)
- Unger J, Chan K, Scovil CY, et al. Intensive balance training for adults with incomplete spinal cord injuries: protocol for an assessor-blinded randomized clinical trial. *Phys Ther*. 2019;99(4):420-427. (protocol)

